# Comparative Analysis of a Large Language Model and Machine Learning Method for Prediction of Hospitalization from Nurse Triage Notes: Implications for Machine Learning-based Resource Management

**DOI:** 10.1101/2023.08.07.23293699

**Authors:** Dhavalkumar Patel, Prem Timsina, Larisa Gorenstein, Benjamin S Glicksberg, Ganesh Raut, Satya Narayan Cheetirala, Fabio Santana, Jules Tamegue, Arash Kia, Eyal Zimlichman, Matthew A. Levin, Robert Freeman, Eyal Klang

## Abstract

Predicting hospitalization from nurse triage notes has significant implications in health informatics. To this end, we compared the performance of the deep-learning transformer-based model, bio-clinical-BERT, with a bag-of-words logistic regression model incorporating term frequency-inverse document frequency (BOW-LR-tf-idf). A retrospective analysis was conducted using data from 1,391,988 Emergency Department patients at the Mount Sinai Health System spanning 2017-2022. The models were trained on four hospitals’ data and externally validated on a fifth. Bio-clinical-BERT achieved higher AUCs (0.82, 0.84, and 0.85) compared to BOW-LR-tf-idf (0.81, 0.83, and 0.84) across training sets of 10,000, 100,000, and ∼1,000,000 patients respectively. Notably, both models proved effective at utilizing triage notes for prediction, despite the modest performance gap. Importantly, our findings suggest that simpler machine learning models like BOW-LR-tf-idf could serve adequately in resource-limited settings. Given the potential implications for patient care and hospital resource management, further exploration of alternative models and techniques is warranted to enhance predictive performance in this critical domain.

## 1. Introduction

Efficient and effective patient triage within the emergency department (ED) plays a pivotal role in enhancing treatment outcomes and optimizing care delivery [1,2,3]. This crucial process involves rapidly identifying patients who require immediate hospitalization upon their arrival. Making the right call can be crucial for patients’ prognosis. One of the pivotal resources for making these critical predictions are nurse triage notes, which provide a wealth of in-depth information about the patient’s condition at presentation [4,5]. In the field of healthcare, machine learning has opened up new avenues for potential improvement in such complex classification tasks, thereby augmenting clinical decision-making processes [6,7]. The recent developments in deep learning and natural language processing (NLP) techniques have further broadened this potential, bringing forth a whole new realm of possibilities for enhancing medical decision-making capabilities.

Among these advanced technological algorithms is the Bidirectional Encoder Representations from Transformers (BERT) model. This model has emerged as a powerful tool in the sphere of NLP [8]. Its excellent performance in numerous NLP tasks [9] has inspired the development of more specialized versions tailored to particular fields, such as the Bio-Clinical-BERT, which was specifically designed to cater to the biomedical field [10]. The focus of this study is to delve into the potential of a fine-tuned Bio-Clinical-BERT model and compare it against a simpler, robust, and more traditional approach, namely the Bag of Words (BOW) Logistic Regression (LR) complemented by the term frequency-inverse document frequency (Tf-Idf) method. The primary objective of our research is to gauge the efficacy of these two methods in predicting hospital admissions using nurse triage notes.

While it’s true that Bio-Clinical-BERT could potentially offer improved accuracy in its predictions, it should be noted that it also requires a substantial investment in terms of computational resources. It necessitates the use of specialized hardware and demands a certain level of software expertise to operate effectively. On the other hand, the LR model paired with the Tf-Idf method, is more resource-efficient and enjoys wide acceptance in the field of text classification due to its simplicity and effectiveness.

Considering these aspects, we have formulated a hypothesis for our study. We hypothesize that the Bio-Clinical-BERT model may surpass the performance of the BOW LR model combined with the Tf-Idf approach in the task of predicting triage outcomes. However, we also speculate that the incremental gains in performance might not necessarily justify the additional demands imposed by the large deep-learning model in terms of computational resources and technical know-how. To test this hypothesis, we have undertaken an extensive study using over one million nurse triage notes collected from a large health system, subjecting both models to the same data for a fair comparison.

The fundamental contribution of this paper is a comprehensive comparison between these two distinct techniques for predicting hospital admission. Our comparison not only looks at the accuracy of these models, but also weighs the trade-offs between predictive accuracy and computational efficiency, a consideration that is often overlooked but is of prime importance in real-world settings – when implementing models. Our aim is to equip healthcare practitioners, researchers, and decision-makers with insights that could potentially aid in enhancing hospital resource management and improve the quality of patient care.

## 2. Methods

### 2.1. Data Sources and Study Design

For the construction and testing of our models, we utilized an extensive dataset from the Mount Sinai Health System (MSHS). This is a diverse healthcare provider based in New York City. In this study, the dataset included Emergency Department (ED) records spanning a five-year period from 2017 to 2022. This dataset was meticulously gathered from five different MSHS hospitals, covering a broad range of population groups and diverse urban health settings.

These five participating hospitals provided a rich source of data for our study, representing different communities in New York City. The hospitals include Mount Sinai Hospital (MSH), a healthcare institution located in East Harlem, Manhattan; Mount Sinai Morningside (MSM), situated in Morningside Heights, Manhattan; Mount Sinai West (MSW), operating in Midtown West, Manhattan; Mount Sinai Brooklyn (MSB), a community-focused health facility located in Midwood, Brooklyn; and Mount Sinai Queens (MSQ) based in Astoria, Queens. The dataset used for our study was compiled using the Epic Electronic Health Records (EHR) software, a tool that aids in efficient data collection, management, and analysis. The dataset was made available by the diligent work of the MSH Clinical Data Science team.

### 2.2. Model Development and Evaluation

In the development and testing of our models, we leveraged data from four hospitals for training, validation, and hyperparameter tuning processes. We elected to use a distinct dataset from Mount Sinai Queens (MSQ) for external testing to ensure our model’s generalizability.

The internal training and validation cohort underwent a rigorous procedure involving five-fold cross-validation. Each fold contained 10,000 records, which were used for hyperparameter tuning. For the external dataset, we experimented with training sets of varying sizes: 10,000, 100,000, and roughly 1,000,000 patients, which represent the complete four-hospital cohort. Subsequently, testing was carried out on 20% of these cohorts’ sizes, taken from the MSQ hospital cohort.

Our study involved two prominent models: bio-clinical-BERT and bag-of-words (BOW) logistic regression models, utilizing term frequency-inverse document frequency (tf-idf) features. These models were employed to predict hospitalization outcomes from nurse triage notes. For bio-clinical-BERT, we adhered to text preprocessing and tokenization guidelines as outlined on the Huggingface.com website [21]. Further details on hyperparameter selection are elucidated in section 3.2 of the results. For BOW LR Tf-Idf, we followed similar methodology outlined in our previous publication [11], covering both text preprocessing and hyperparameter selection processes.

BERT: BERT, or Bidirectional Encoder Representations from Transformers, is a model designed for natural language processing tasks. It learns from the context of both preceding and following words, making it “bidirectional”. This feature sets BERT apart, as it allows for a better understanding of language semantics. This model is pre-trained on large corpora and can be fine-tuned for specific tasks.

Bag of Words (BOW): The Bag of Words model is a simple technique in natural language processing. It represents text data by counting the frequency of each word, disregarding the order in which they appear. Each unique word forms a feature, and the frequency of the word represents the value of that feature. However, this method can overlook context and semantics due to its simplicity.

TF-IDF: TF-IDF stands for Term Frequency-Inverse Document Frequency. It’s a numerical statistic that reflects how important a word is to a document in a collection. It is a combination of two metrics: Term Frequency, which is the number of times a word appears in a document, and Inverse Document Frequency, which diminishes the weight of common words and amplifies the weight of rare words across the entire dataset. This helps in reducing the impact of frequently used words and highlights more meaningful terms.

### 2.3. Study Population

The demographic for this study included adult patients aged 18 years and above. These were patients who made ED visits within the specified five-year period of 2017-2022 across the five participating MSHS hospitals.

### 2.4. Outcome Definition

The primary outcome for our study was to ascertain our models’ effectiveness in predicting hospitalization. This prediction was based on two main types of data: tabular Electronic Health Records (EHR) and nurse triage notes.

### 2.5. Model Evaluation and Comparison

To rigorously assess the performance of our models, we utilized various metrics such as Area Under the Receiver Operating Characteristic curve (AUC), sensitivity, specificity, and precision. These metrics allowed us to thoroughly evaluate the bio-clinical-BERT [10] and BOW logistic regression models with tf-idf features, and compare their capabilities in predicting hospitalization from nurse triage notes.

### 2.7. Ethical Considerations

This study, being retrospective in nature, was reviewed and approved by an ethical institutional review board (IRB) committee from MSHS. The IRB committee deemed that due to the retrospective nature of the study, the requirement for informed consent was waived.

### 2.8. Statistical Analysis

Our statistical analyses were conducted using Python (Version 3.9.12). We presented continuous variables as median (IQR) and categorical variables as percentages for better interpretability. To identify words linked to hospital admission within nurse triage notes, we calculated the Odds Ratio (OR) and Mutual Information (MI) [11]. Statistical tests such as the χ2 test and Student’s t-test were employed for comparing categorical and continuous variables, respectively. A p-value of less than 0.05 was considered statistically significant. For evaluating our models, Receiver Operating Characteristic (ROC) curves were plotted, and metrics including AUC, sensitivity (recall), specificity, and positive predictive value (precision) were derived, with Youden’s index used to determine optimal cut-off values.

### 2.9 Technical Architecture

The technical experiments involved in this study were conducted within a controlled hospital infrastructure that used an On-Premises Centos Linux environment in conjunction with Azure Cloud infrastructure. For the BOW TF-IDF experiments, we elected to use the Centos Linux OS. In contrast, the BERT experiment was conducted using a Standard_NC6 GPU instance on Azure Cloud. This instance came with 1 16GB GPU, 6 vCPU, and incurred a cost of approximately $80 during the training phase. **Figure 1** offers a detailed depiction of the fundamental technical architecture employed for training the BERT and LR-TFID models, across multiple patient datasets.

**Figure 1.**
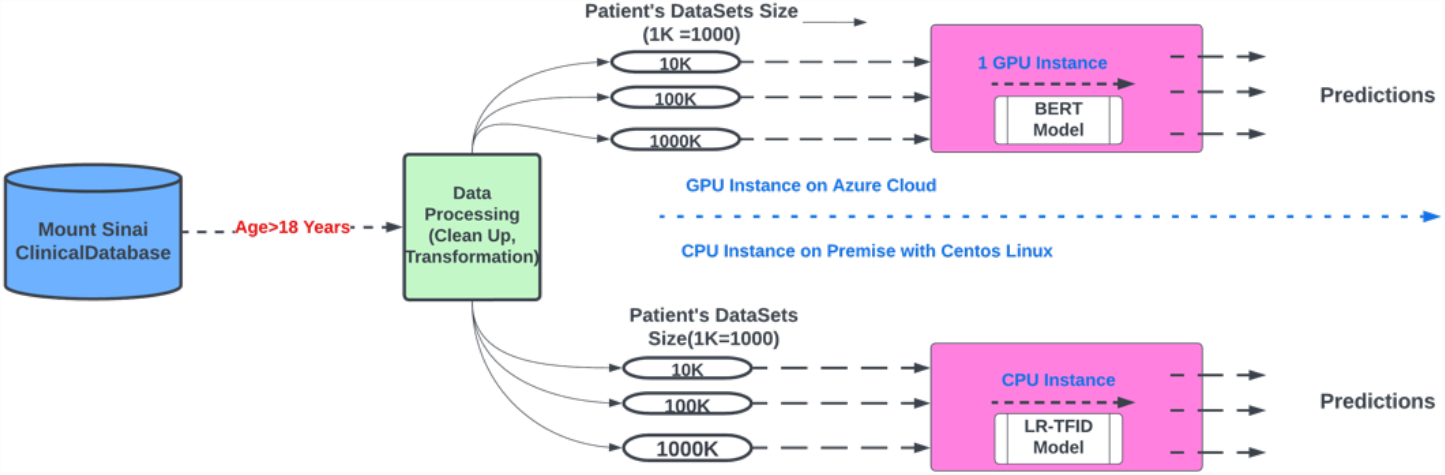
illustrates the process flow of multiple patient datasets passing through two different models with GPU and non-GPU instances.

## 3. Results

### 3.1. Patient Population and Data

Our study incorporated data from 1,745,199 patients drawn from the Mount Sinai Health System. Upon exclusion of patients below 18, we had 1,391,988 participants in the study. These patients visited the ED between 2017 and 2022. **Table 1** presents a summary of the patient characteristics.

**Table 1:** Demographic distribution in the research cohorts (Abbreviations: IQR – interquartile range, MSQ – Mount Sinai Queens)

|  | 4 hospitals<br>Median<br>(IQR) | MSQ<br>Median<br>(IQR) | P value |
| --- | --- | --- | --- |
| **Total number of patients** | 1,391,988 (includes MSH, MSM, MSW, MSB and MSQ Facilities) |  |  |
| **Age** | 48.0<br>(32.0 – 75.0) | 45.0 (30.0 – 75.0) | P<0.001 |
| **Sex** | Female:<br>52.8% | Female:<br>50.1% | P<0.001 |
|  | Male:<br>47.20% | Male:<br>49.9% |  |
| **Race:** |  |  | P<0.001 |
| **Black** | 382,898<br>(34.50%) | 45,696<br>(16.22%) |  |
| **White** | 265,457<br>(23.92%) | 77,622<br>(27.56%) |  |
| **Other** | 461,917<br>(41.58%) | 158,398<br>(56.22%) |  |

The median number of words per triage note was 19.0 (IQR 12.0 – 31.0). Top ten words associated with the highest MI score regarding hospital admission are outlined in **Table 2**.

**Table 2:** Odds Ratios and Mutual Information values for words linked to admission to hospital wards, sorted by highest Mutual Information values. Abbreviations: OR – Odds Ratio, MI – Mutual Information.

| Word | OR for Admission | MI for Admission | p value |
| --- | --- | --- | --- |
| Sent | 3.6 | 16.4 | <0.001 |
| Pt | 1.6 | 15.8 | <0.001 |
| Per | 2.3 | 15 | <0.001 |
| Of | 1.3 | 12.7 | <0.001 |
| Home | 2.2 | 11.5 | <0.001 |
| EMS | 2.2 | 10.8 | <0.001 |
| Weakness | 3.6 | 10.8 | <0.001 |
| Chest | 1.4 | 8.9 | <0.001 |
| SOB | 2.1 | 8.8 | <0.001 |
| BIBA | 2.1 | 7.9 | <0.001 |

### 3.2. Hyperparameter Tuning Results

A comprehensive hyperparameter tuning process was performed. The best hyperparameters were identified for each model based on their performance during the five-fold cross-validation on the training validation set. The results of the BERT hyperparameter tuning process can be found in **Table 3**.

**Table 3:** Hyperparameter tuning in the internal training/validation cohorts using five-fold experiments (Abbreviations: BS: Batch Size, LR: Learning Rate, ML: Max Length)

|  | LR:2e<sup>-5</sup><br>Epoch: 3 |  |  |  | LR:3e<sup>-5</sup><br>Epoch: 3 |  |  |  | LR:5e<sup>-5</sup><br>Epoch: 3 |  |  |  |
| --- | --- | --- | --- | --- | --- | --- | --- | --- | --- | --- | --- | --- |
|  | BS: 64 | BS:128 | BS: 128<br>ML:128 | BS:256<br>ML: 64 | BS: 64 | BS:128 | BS:128<br>ML:128 | BS:256<br>ML:64 | BS: 64 | BS:128 | BS:128<br>ML:128 | BS:256<br>ML:64 |
| BERT | 0.78±SD | 0.80±SD | 0.80±SD | 0.79±SD | 0.79±SD | 0.79±SD | 0.78±SD | 0.78±SD | 0.79±SD | 0.80±SD | 0.79±SD | 0.79±SD |

### 3.3. Model Performance

After training the bio-clinical-BERT and LR-tf-idf models on the four hospitals’ data, we evaluated their performance on the held-out test data from Mount Sinai Queens (MSQ). The area under the receiver operating characteristic (AUC) values were calculated for each model. The bio-clinical-BERT model achieved AUCs of 0.82, 0.84, 0.85 while the LR-tf-idf model had AUCs of 0.81, 0.83, 0.84 for training on 10,000, 100,000, and ∼1,000,000 patients.

**Figure 1** shows the ROC and AUC comparisons between the two models. The bio-clinical-BERT model consistently outperformed the LR-tf-idf model in terms of AUC across the different training set sizes (10,000, 100,000, and ∼1,000,000 patients), albeit by a small margin.

**Figure 1:**
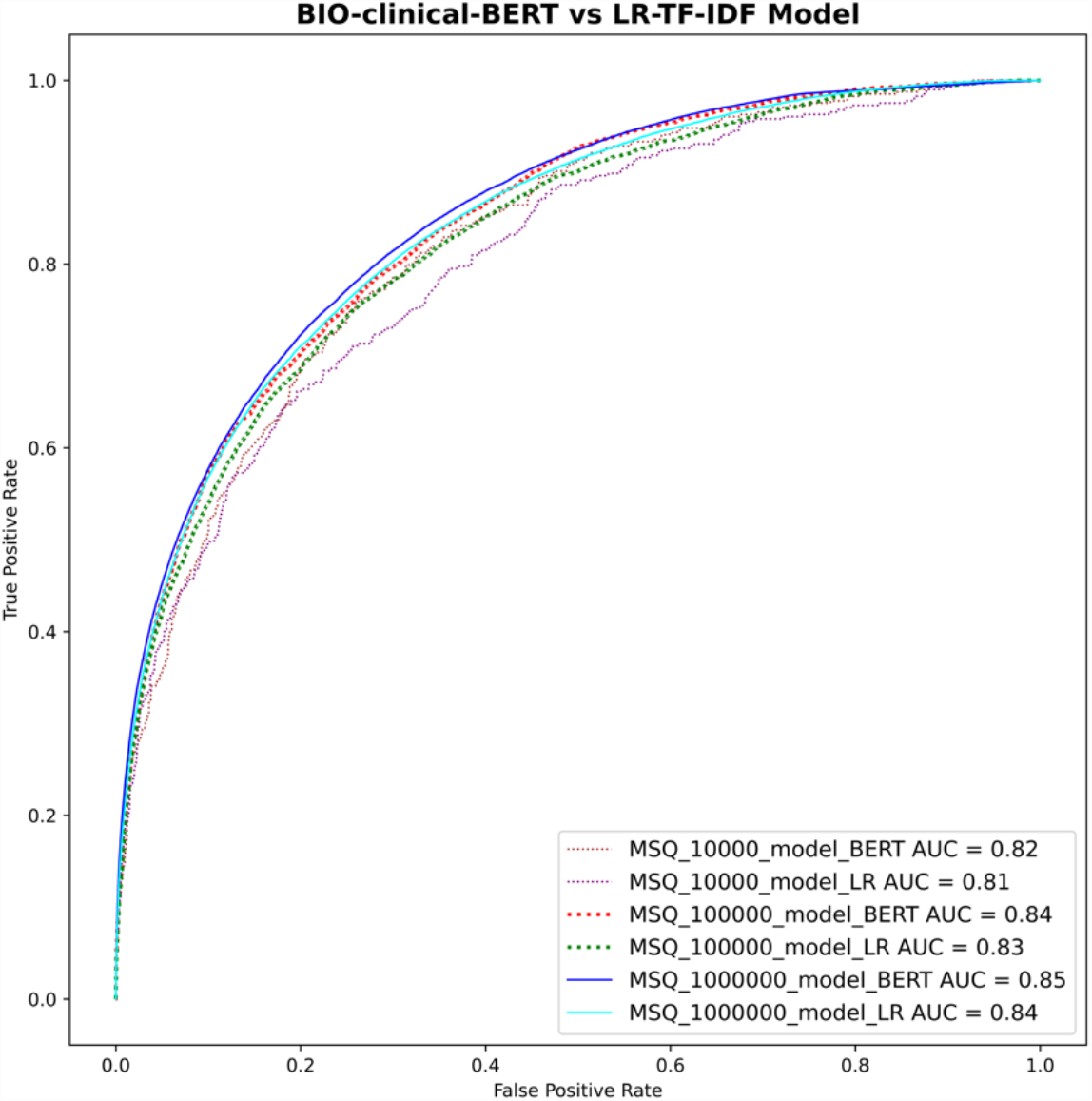
receiver operating characteristic curves (ROC) of the two models tested on increasing training sample sizes.

In addition to the AUC comparisons, we also calculated other performance metrics, such as sensitivity, specificity, and precision, for both models (**Table 4 and Table 5**). The probability cut-off values for these metrics were calculated using Youden’s index. These results further demonstrated the superior performance of the bio-clinical-BERT model compared to the LR-tf-idf model.

**Table 4:** metrics results for the training/testing (external) cohort for bio-clinical-BERT (Abbreviations: AUC – Area under the curve)

| Training Data Size | AUC_Score | Sensitivity | Specificity | Precision | F1_Score |
| --- | --- | --- | --- | --- | --- |
| 10000 | 0.82 | 0.76 | 0.74 | 0.36 | 0.49 |
| 100000 | 0.84 | 0.74 | 0.77 | 0.39 | 0.51 |
| 1000000 | 0.85 | 0.39 | 0.96 | 0.67 | 0.50 |

**Table 5:** metrics results for the training/testing (external) cohort for tf-idf-LR (Abbreviations: AUC – area under the curve)

| Training Data Size | AUC_Score | Sensitivity | Specificity | Precision | F1_Score |
| --- | --- | --- | --- | --- | --- |
| 10000 | 0.81 | 0.66 | 0.80 | 0.40 | 0.50 |
| 100000 | 0.83 | 0.75 | 0.74 | 0.37 | 0.50 |
| 1000000 | 0.84 | 0.71 | 0.80 | 0.42 | 0.53 |

## 4. Discussion

In this study, we sought to compare the performance of two predictive models, bio-clinical-BERT and LR-tf-idf, in predicting hospitalizations based on nurse triage notes. The findings of our study suggest that while bio-clinical-BERT does marginally outperform LR-tf-idf in this predictive task, the difference in their performance is relatively minor.

Such results echo the findings of previous studies in the field, which have often found BERT-based models to have a slight edge over traditional machine learning methods like LR-tf-idf in various natural language processing tasks [12, 13]. However, it’s essential to note that the marginal difference observed in our study suggests that, given certain limitations such as constraints on hardware, software expertise, or budget, hospitals might lean towards simpler machine learning methods. The rationale behind such a choice would lie in the ease of implementing these simpler methods, as well as their relatively less demanding computational requirements.

The comparison of different models in the biomedical domain has been the focus of numerous previous studies. For instance, Chen et al. conducted a rigorous assessment of transformer-based ChatGPT models in tasks like reasoning and classification [14]. Their study found that fine-tuning remained the most effective approach for two central NLP tasks. However, it’s interesting to note that the basic Bag-of-Words model managed to deliver comparable results to the more complex Language Model prompting. It should be noted that the creation of effective prompts required a substantial resource investment.

In another study, Xavier et al. compared three different model types for a multiclass text classification task, which involved the assignment of protocols for abdominal imaging CT scans [15]. These models spanned a range from conventional machine learning and deep learning to automated machine learning builder workflows. While the automated machine learning builder boasted the best performance with an F1 score of 0.85 on an unbalanced dataset, the Tree Ensemble machine learning algorithm was superior on a balanced dataset, delivering an F1 score of 0.80.

A further study delved into the evaluation of Machine Learning multiclass classification algorithms’ performance in classifying proximal humeral fractures using radiology text data [16]. Several statistical ML algorithms performed reasonably, with a BERT model showcasing the best accuracy of 61%. In another relevant study conducted by Ji et al., various models pretrained with BERT were compared for medical code assignment based on clinical notes. Interestingly, it was found that simpler artificial neural networks could sometimes outperform BERT in certain scenarios [17]. This study, among others, offers further support to our recommendation for hospitals with limited resources to consider simpler, less resource-demanding methods for achieving comparable predictive performance.

In the specific task of predicting hospitalization, both methods in our study effectively leveraged the rich information found within nurse triage notes. This finding aligns with those from other studies [18, 19, 20]. For instance, a study by Zhang et al. that evaluated logistic regression and neural network modeling approaches in predicting hospital admission or transfer after initial ED triage presentation found that the patient’s free text data regarding referral improved overall predictive accuracy [18]. Similarly, Raita et al. utilized machine learning models to predict ED outcomes and demonstrated superior performance in predicting hospitalization [19].

The results of our study carry practical implications for healthcare organizations. The ability to predict hospitalization from nurse triage notes could lead to significant improvements in patient care by facilitating efficient resource allocation, optimizing bed management, and improving patient flow. The choice between the use of bio-clinical-BERT and simpler methods, such as LR-tf-idf, should be influenced by the specific context of the organization, including factors such as available computational resources, software expertise, and desired model performance.

Our study is not without limitations. For instance, the data used for our study is specific to MSHS hospitals, which might not be representative of other healthcare systems, potentially limiting the generalizability of our findings. Despite using multi-site data, representing the diverse NY city population, and an external validation site for our final analysis, we acknowledge the need for further studies with more diverse datasets. We also recognize that we did not explore the potential of combining both methods or other potential techniques that could enhance these models’ performance. Moreover, the field of NLP is advancing fast, with new large language models constantly evolving.

Future research could focus on the exploration of BERT models that are pre-trained and trained from scratch on a site’s entire textual data. Although such an approach may demand significant resources and be computationally intensive, it might yield better performance by capturing the unique characteristics and language patterns of a specific healthcare setting. The exploration of other pre-trained language models or more advanced natural language processing techniques could also pave the way for the development of more effective hospitalization prediction methods based on nurse triage notes.

In conclusion, our study demonstrates that while bio-clinical-BERT does marginally outperform LR-tf-idf in predicting hospitalization from nurse triage notes, the difference is small enough to suggest that simpler methods might be viable for hospitals with limited resources. More research is needed to identify alternative methods that can enhance these models’ performance in predicting hospitalization, ultimately improving patient care and hospital resource management.

Through a rigorous investigation of bio-clinical-BERT and LR-tf-idf models’ performance, our study contributes to the growing body of literature in the field of natural language processing and machine learning in healthcare. It emphasizes the importance of considering the trade-offs between model complexity and performance when deploying predictive tools in clinical settings, highlighting that sometimes, simpler methods can prove almost as effective as more complex ones.

## Data Availability

All data produced in the present study are available upon reasonable request to the authors

